# Cost effectiveness of interventions for Post-Traumatic Stress Disorders following major incidents including terrorism and pandemics

**DOI:** 10.1101/2020.06.26.20141051

**Authors:** Nicole Hogan, Martin Knapp, David McDaid, Mark Davies, Chris R. Brewin

## Abstract

**Background:** Post-traumatic stress disorder (PTSD) is commonly experienced in the aftermath of major incidents such as terrorism and pandemics. Well-established principles of response include effective and scalable treatment for individuals affected by PTSD. In England, such responses have combined proactive outreach, screening, and evidence-based interventions (a “screen-and-treat” approach), but little is known about the cost-effectiveness of this approach.

**Methods:** A decision modelling analysis was undertaken to estimate the costs per quality adjusted life year (QALY) gained from a screen-and-treat approach compared to treatment-as-usual. Model input variables were drawn from relevant empirical studies in the context of terrorism and the unit costs of health and social care in England. The model was run over a five-year time horizon for a hypothetical cohort of 1,000 exposed adults from the perspective of the National Health Service and Personal Social Services in England.

**Results:** The incremental cost per QALY gained was £8,297. This would be considered cost-effective 95% of the time at a willingness-to-pay threshold of £20,000 per QALY gained, the threshold associated with NICE. Sensitivity analysis confirmed this result was robust.

**Conclusions:** A screen-and-treat approach for identifying and treating PTSD in adults following major incidents appears cost-effective in England compared to treatment-as-usual through conventional primary care routes. This finding was in the context of terrorism but can be translatable into other major-incident related scenarios including the current COVID-19 pandemic in lieu of data on the impact of this pandemic.

**Competing interest statement:** The authors have declared no competing interest.

**Funding statement:** No additional funding was provided for this study

**Author Declarations:** All relevant ethical guidelines have been followed; any necessary IRB and/or ethics committee approvals have been obtained and details of the IRB/oversight body are included in the manuscript.

Yes

All necessary patient/participant consent has been obtained and the appropriate institutional forms have been archived.

Yes

I understand that all clinical trials and any other prospective interventional studies must be registered with an ICMJE-approved registry, such as ClinicalTrials.gov. I confirm that any such study reported in the manuscript has been registered and the trial registration ID is provided (note: if posting a prospective study registered retrospectively, please provide a statement in the trial ID field explaining why the study was not registered in advance).

Yes

I have followed all appropriate research reporting guidelines and uploaded the relevant EQUATOR Network research reporting checklist(s) and other pertinent material as supplementary files, if applicable.

Yes

## Background

Posttraumatic stress disorder (PTSD), a severe and chronic condition associated with high levels of functional impairment, is the most common single psychiatric outcome of major incidents worldwide, including pandemics (Bonanno, Brewin, Kaniasty, & La Greca, 2010; Ghebreyesus, 2020; Mak et al., 2009; Maunder et al., 2006). Following exposure to a terrorist attack, for example, adults have a prevalence of PTSD estimated at 30-40% (Whalley & Brewin, 2007). Other sequelae include anxiety, depression, and substance misuse. Despite the existence of effective psychological therapies, PTSD typically remains untreated, whether or not it occurs in the context of a major incident (Pfefferbaum et al., 2002; Wang et al., 2005; WTC Medical Working Group, 2008). The reasons for this are well-understood: a combination of low priority at strategic planning and policy development level, general low levels of public understanding, poor recognition in primary care, and the avoidance that is one of the defining symptoms. In recent years, health systems have begun to address this problem by instituting proactive outreach to affected populations, often coupled with screening and signposting into evidence-based treatments (Brewin et al., 2008; Dyb et al., 2014; French et al., 2019; Maslow et al., 2015). In this paper we report the first systematic attempt to assess the cost-effectiveness of this approach. Data are taken from studies of terrorist attacks affecting UK residents, but our conclusions are relevant to other major incidents including pandemics and can inform responses to COVID-19.

It is important to highlight upfront that psychological reactions to trauma vary in severity and duration. Fortunately, the majority of individuals directly exposed to trauma will not develop PTSD as many symptoms of distress will naturally decline and are unlikely to have long-term implications (Whalley & Brewin, 2007). A sizeable minority require mental health services and a small proportion need long-term intervention.

In England, the public health approach following recent major incidents has been to institute an initial period of “watchful waiting”, which for the majority will allow natural coping resources and social support to lead to spontaneous remission, and mitigates the deployment of resources on individuals with transient conditions. After this initial delay outreach and screening are offered to populations at high-risk of developing PTSD. This is done using brief, validated instruments (DoH, 2009; National Institute for Health and Care Excellence (NICE), 2018a). A stepped model of care is recommended, which starts with assessment and facilitates access to mental health services if immediate and short-term distress does not resolve. Trauma-focussed cognitive behavioural therapy (CBT) is a highly-effective treatment for PTSD, typically delivered over 8 to 12 sessions (NICE, 2018a). Approaches that link evidence-based treatment with outreach and screening are known as “screen-and-treat” or “outreach-and-screen”.

There have been several bespoke screen-and-treat programmes implemented in England in the context of recent major incidents. These include the Trauma Response Programme following the London bombings, the Screen and Treat Programme following the terrorist attacks in Tunisia, Brussels and Paris, the Manchester Resilience Hub following the Manchester Arena bombing, and the Health and Wellbeing Service after the Grenfell Tower fire in West London. Evaluations of those programmes have concluded that screen-and-treat approaches may be a clinically effective approach for addressing mental health needs following major incidents (Brewin et al., 2010a; Cyhlarova, Knapp, & Mays, 2019; French et al., 2019; Gobin et al., 2018; Kerslake Report, 2018).

With these screen-and-treat approaches, evidence on cost-effectiveness as well as clinical effectiveness is vital to support decisions on both short- and longer-term resourcing. The aim of our study is to use decision analytic modelling to evaluate the cost-effectiveness of a screen-and-treat approach for identifying and treating PTSD following major incidents compared to treatment-as-usual delivered as a result of identification through conventional primary care routes.

## Methods

### Study design and assumptions

Economic modelling can be used to synthesise evidence on the effectiveness of different interventions with evidence on the costs and consequences of these actions. The approach is used as standard by the National Institute for Health and Care Excellence in England (NICE) when looking at the case for investing in any health care intervention. Assumptions in models can be varied to help provide decision makers with a range of policy-relevant information, including uncertainty on evidence of effect, as well as level of uptake and sustained use. Models can also be used to extrapolate longer-term impacts than those seen in many empirical trials.

A decision tree was constructed in Microsoft Excel comparing the screen-and-treat approach with treatment-as-usual for a hypothetical cohort of individuals. Each pathway was assumed to consist of 1,000 adults who were directly exposed to terrorism at time zero. The clinical pathway for the screen-and-treat intervention group was based on a simplified version of the mental health responses implemented following the London bombings in 2005 and 2017. The intervention pathway assumed that individuals were screened with the Trauma Screening Questionnaire (TSQ) at 3 months (Brewin et al., 2002). The intervention pathway is shown in Figure 1. It was not necessary to incorporate Markov cycles into the model as participants were unlikely to move between states after the initial period (other than to the absorbent death state, which would occur at equal rates across the two groups).

**Figure 1.**
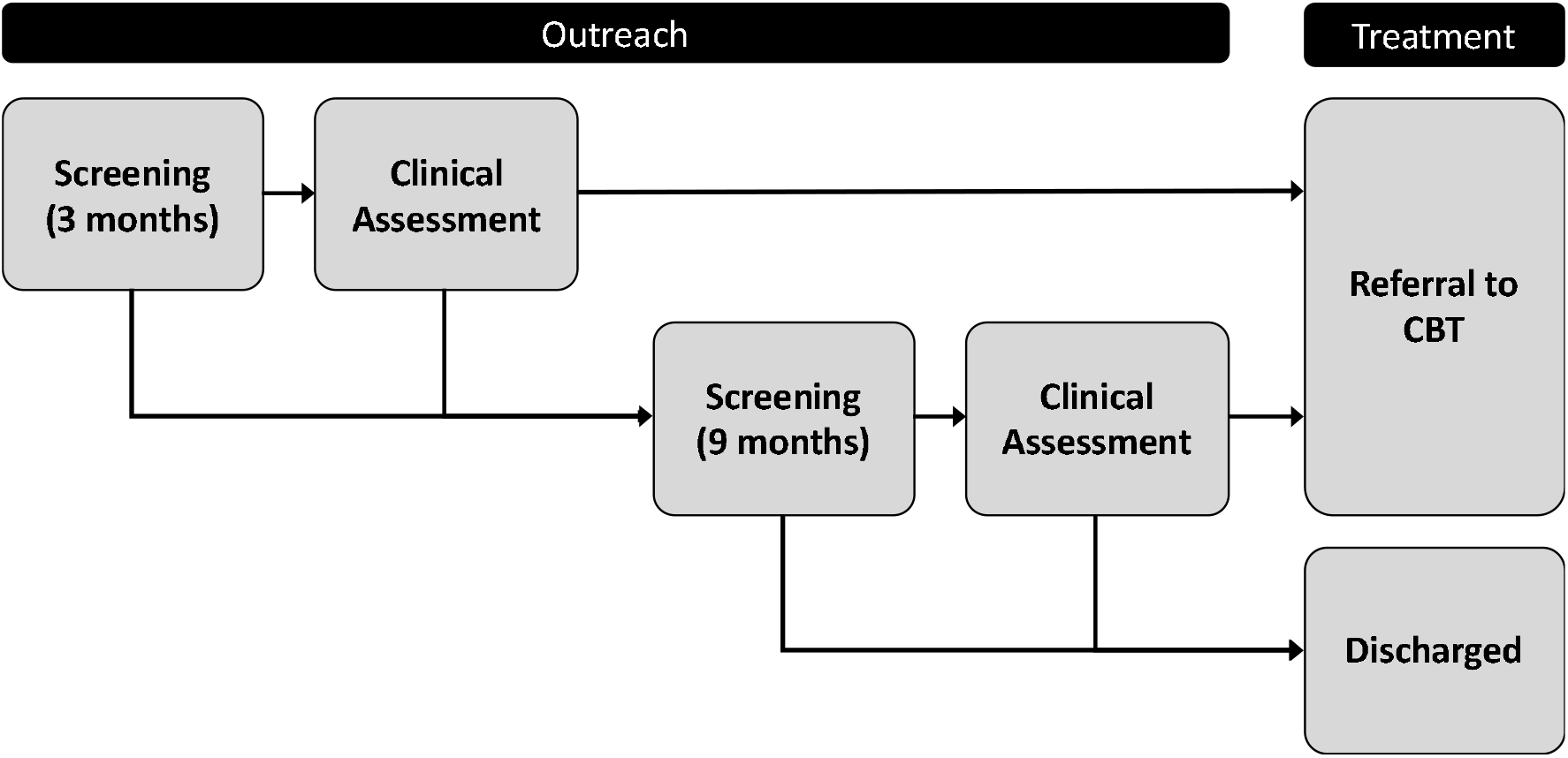
Screen-and-treat clinical pathway.

The participants who screened positive would then have a clinical assessment in the form of the Structured Clinical Interview for DSM-5 (SCID-5) (First, Williams, Karg, & Spitzer, 2015). This is a commonly employed measure and assumed to be 100% accurate. “True positives” would then be referred for CBT through Improving Access to Psychological Therapies (IAPT) services, receiving an average of 12 individual weekly sessions over three months. Rates of uptake and completion of CBT were modelled separately. The “false positives” were screened out as they did not have PTSD. Subsequently at 9 months, all participants in the intervention pathway who had not been referred to treatment were screened again using the TSQ, in an attempt to capture the “false negatives” from the first round of screening as well as those participants that developed delayed-onset PTSD. The process of assessment using the SCID-5 and again referral for treatment with CBT was offered.

The treatment-as-usual comparator group received no treatment, unless the individual was detected as having PTSD through conventional primary care routes. Participants in the comparator pathway with PTSD who were detected in primary care were also referred to IAPT services for treatment. At the end of the clinical pathway, the proportions of participants in the intervention and comparator groups with PTSD, partial PTSD, and no PTSD were calculated. Partial PTSD was a health state intended to reflect those individuals experiencing sub-clinical symptoms.

Costs and outcomes conditional on the individual‘s health state were simulated over a five-year time horizon from a National Health Service (NHS) and Personal Social Services (PSS) perspective to capture the longer-term and wider consequences of PTSD. This is the perspective adopted by NICE in their appraisals. Five years is a plausible time frame as we expect some persistence of effect (Brewin et al., 2010a). All costs were inflated by 1.5% annually to 2018 prices in British Pounds (GBP) and both costs and outcomes were discounted over the five-year time horizon at 3.5% per annum. This discount rate was chosen to reflect central government guidance on appraisal and evaluation (HM Treasury, 2018).

It was conservatively assumed that PTSD was the only mental health impact, and comorbidity with other mental health conditions was not modelled. This simplification was made because of a lack of data to the contrary. In any event, PTSD represents the overwhelming majority of mental health need following terrorism and successful treatment for PTSD also leads to remission of comorbid conditions (Whalley & Brewin, 2007; Brewin et al., 2008; Neria et al., 2008). The model was further simplified to ignore the risk of relapse because gains from CBT tend to be maintained following a single trauma (Brewin et al., 2010a; Santiago et al., 2013).

Incremental cost per quality-adjusted-life-year (QALY) was then calculated. This is the reference case measure used by NICE when assessing the case for investing in health care interventions and enables comparison between investment choices for different health care and public health programmes.

### Model parameters

Model inputs are shown in Table 1. Where possible, the most relevant “local” parameter estimates were used. The prevalence of PTSD in adults was assumed to be 31% based on the proportion of survivors that required treatment for PTSD following the London bombings (Brewin et al., 2010a). This is consistent with an international systematic review finding a mean prevalence of 29·8% in adult victims of terrorism (Garcia-Vera, Sanz, & Gutierrez, 2016). Furthermore, it was assumed that for 85% of participants the onset of PTSD would be “immediate” (occur within three months) and the remaining 15% would have a “delayed” onset (at six months). This is consistent with evidence on the prevalence of delayed-onset PTSD (Andrews, Brewin, Philpott, & Stewart, 2007).

**Table 1.** Model inputs

| Description | Abbreviation | Reference Case | Standard Error | Reference |
| --- | --- | --- | --- | --- |
| **Probabilities** |  |  |  |  |
| Prevalence of PTSD | prev_PTSD | 0.310 | 0.01 | Brewin et al., 2010a |
| Proportion of immediate-onset PTSD | prop_immedPTSD | 0.850 | - | Andrews et al., 2007 |
| Proportion of delayed-onset PTSD | prop_delayPTSD | 0.150 | - | Andrews et al., 2007 |
| Sensitivity of TSQ (3-months) | sens_TQS3 | 0.948 | 0.01 | Brewin et al., 2010b |
| Specificity of TSQ (3-months) | spec_TSQ3 | 0.255 | 0.01 | Brewin et al., 2010b |
| Sensitivity of TSQ (9-months) | sens_TSQ9 | 0.872 | 0.01 | Brewin et al., 2010b |
| Specificity of TSQ (9-months) | spec_TSQ9 | 0.393 | 0.01 | Brewin et al., 2010b |
| Detection of PTSD through primary care routes | p_detectPTSD | 0.050 | 0.01 | Ehlers et al., 2009; Rubin & Wessely, 2013 |
| CBT uptake | p_CBTuptake | 0.875 | 0.01 | Brewin et al., 2010a |
| CBT completion | p_CBTcomplete | 0.871 | 0.01 | Brewin et al., 2010a |
| Spontaneous remission | p_sponremit | 0.200 | 0.01 | Authors' assumption; Santiago et al., 2013 |
| Recovery with CBT | p_CBTrecover | 0.624 | 0.01 | Brewin et al., 2010a |
| Partial recovery with CBT | P_CBTpartrecover | 0.136 | 0.01 | Authors' assumption; Brewin et al., 2010a |
| **Costs (GBPs at 2018 prices)** |  |  |  |  |
| PTSD | c_PTSD | 1173 | 235 | NICE, 2018b |
| Partial PTSD | c_partPTSD | 642 | 128 | Authors' assumption; NICE, 2018b |
| No PTSD | c_noPTSD | 110 | 22 | NICE, 2018b |
| Identifying and screening | c_TSQ | 588 | 118 | Fuchkan, 2015 |
| Clinical assessment | c_SCID | 588 | 118 | Fuchkan, 2015 |
| CBT IAPT treatment | c_IAPT | 1,710 | 342 | Curtis & Burns, 2018 |
| **Utilities (QALYs)** |  |  |  |  |
| PTSD | u_PTSD | 0.610 | 0.05 | Gospodarevskaya & Segal, 2015 |
| Partial PTSD | u_partPTSD | 0.730 | 0.05 | Authors' assumption; Gospodarevskaya & Segal, 2015 |
| No PTSD | u_noPTSD | 0.850 | 0.05 | Gospodarevskaya & Segal, 2015 |

Studies analysing data from the London bombings were used to estimate the specificity and sensitivity of the TSQ at 3-months and 9-months (Brewin, Fuchkan, & Huntley, 2010b) and the probabilities of uptake of CBT, completion of CBT, and recovery or partial recovery with CBT (Brewin et al., 2010a). The probability of detection of PTSD in primary care was approximated at 5% based on studies on PTSD detection rates in conventional primary care settings in England (Ehlers, Gene-Cos, & Perrin, 2009), supported by repeated reports of lack of referral of survivors of terrorist attacks by family doctors (Brewin et al., 2010a; Cylharova et al., 2019).

The probability of spontaneous remission was based on findings from a recent systematic review that 34·8% of those with PTSD following exposure to intentional trauma remit after 3 months (Santiago et al., 2013). However, that figure was lowered at the discretion of the authors to 20%. This was for three key reasons. First, individuals who judge they are likely to get better on their own may not engage with the programme, while those who do engage may have a more chronic course. Second, remission is based on no longer meeting diagnostic criteria for PTSD and in practice many people may have significant residual symptoms that could benefit from treatment. For example, they may still have clinical levels of depression or have a phobic condition. Third, remission in the literature has only been measured at one point in time and people may fluctuate between meeting and not meeting diagnostic criteria if they are followed up for longer. For example, they might have another onset triggered by an inquest or court case.

The costs per person for screening and assessment, both averaging £588, were calculated using 2015 data from the London bombings inflated to 2018 prices (Fuchkan, 2015). These costs included start-up and management of the programme, finding, screening and assessing participants, and referral management. The cost of treatment through IAPT (£1,710) was calculated using unit costs of health and social care at 2018 prices (£95) multiplied by an average number of 12 × 1.5 hour sessions (Curtis & Burns, 2018).

The costs of medication and health and personal social services were approximations based on a recent expert panel review convened in England to review the economic evidence on PTSD (NICE, 2018b). The annual costs of being in the PTSD or no PTSD health states were approximated by the panel as £1,173 and £110 per person respectively at 2018 prices. Forecast costs included medication, inpatient hospital stays, outpatient visits, general practitioners and district nursing, outreach and home help, and psychological treatment. The cost of being in the partial PTSD health state was assumed to be mid-way between having PTSD and not having PTSD (£642) to capture the associated costs of sub-clinical symptoms. Our analysis is conservative as productivity losses arising from disrupted education or employment were excluded given the NHS and PSS perspective taken.

QALYs were used as the measurement of health gain. A preference-weight of “1” equates to perfect health, “0” to death, and negative values (worse than death) are permitted. We were unable to identify UK utility values for our study population, and instead the PTSD and no PTSD utilities, 0·610 and 0·850 respectively, were based on an Australian study in the context of sexual abuse (Gospodarevskaya & Segal, 2015). The utility value for sub-clinical PTSD resulting from partial recovery with CBT was approximated as the mid-point between having PTSD and not having PTSD.

### Sensitivity analysis

One-way sensitivity analysis was conducted to test the robustness of the results by using standard errors to vary the input variables one at a time on a 95% confidence interval, assuming a normal distribution. Standard errors were approximated as 0·01 for probabilities (to reflect minimal uncertainty), as 0·05 for outcomes (to reflect reasonable upper and lower values that did not overlap between health states), and as 20% of the deterministic input for costs (to reflect the potential for considerable variation).

A probabilistic sensitivity analysis with 1,000 iterations was also conducted to vary input variables simultaneously. A gamma distribution was assumed for costs because the data are skewed and constrained between zero and positive infinity, and a beta distribution was assumed for probabilities and outcomes because the data are binomial and constrained between zero and one.

## Results

### Cost-effectiveness

The incremental cost-effectiveness ratio (ICER) was defined in the conventional way as:

ICER=(Δ Costs)/(Δ Effects)

The total costs and effects for 1,000 individuals and the average costs and effects per person are shown as accrued over the five-year time horizon (Table 2). The ICER (£8,972) was expressed as the incremental cost per QALY gained compared to the comparator. NICE in England employs guidelines that compare ICER values to a willingness-to-pay threshold of £20,000 per QALY. Given the ICER is below that threshold, a screen-and-treat approach would be considered a cost-effective option based on the model inputs.

**Table 2.**
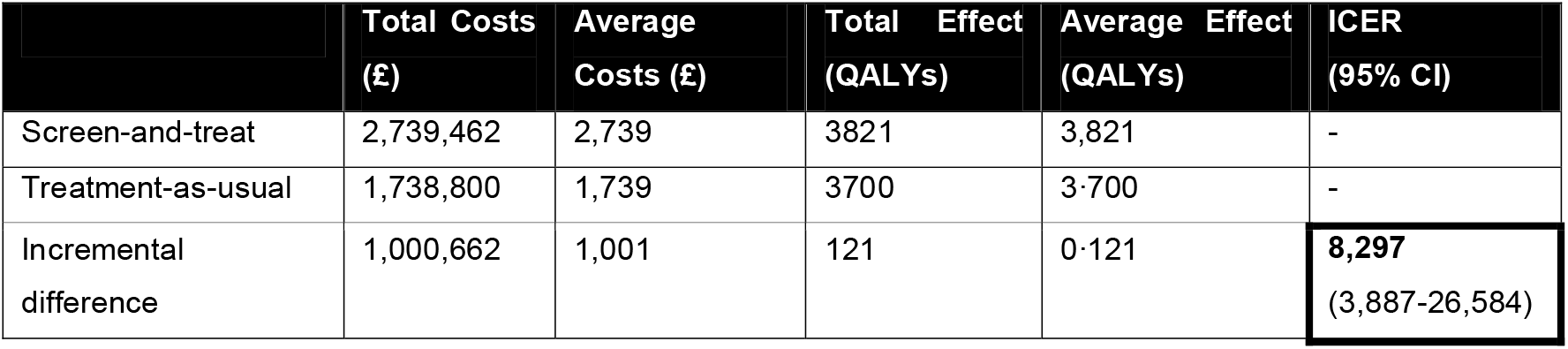
Incremental cost-effectiveness ratio (ICER)

### One-way sensitivity analysis

The sensitivity of the incremental cost-effectiveness ratio to an increase or decrease in model input variables is shown in the tornado diagram (Figure 2). In short, the three key drivers of the ICER were the utility of being in the PTSD or no PTSD health states (u_PTSD, u_noPTSD) and the costs associated with having PTSD (c_PTSD).

**Figure 2.**
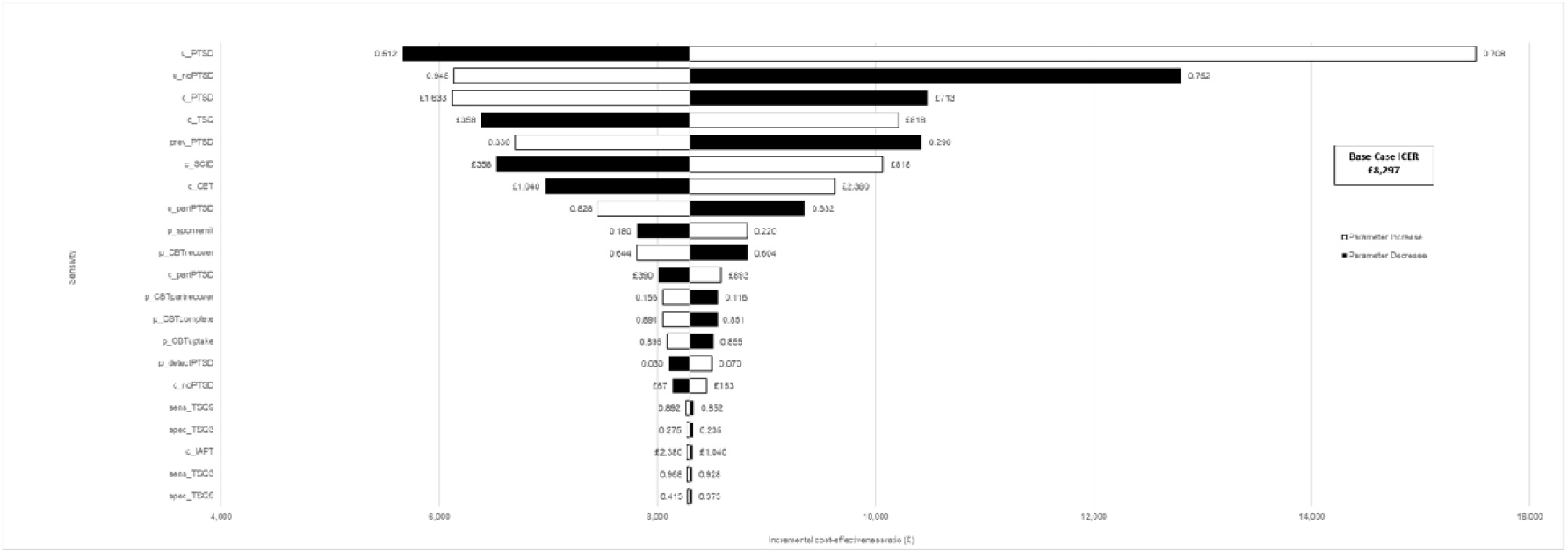
Tornado diagram.

### Probabilistic sensitivity analysis

The incremental costs and benefits, having run the model over 1,000 iterations, are presented as a cost-effectiveness acceptability curve (Figure 3). This curve represents the proportion of iterations (out of 1,000) that would be considered cost-effective at various willingness-to-pay thresholds. The screen-and-treat approach was found to have a 95% chance of being considered cost-effective given a willingness-to-pay threshold of £20,000 per QALY. The probabilistic sensitivity analysis was also used to calculate the 95% confidence interval (£3,887 to £26,584).

**Figure 3.**
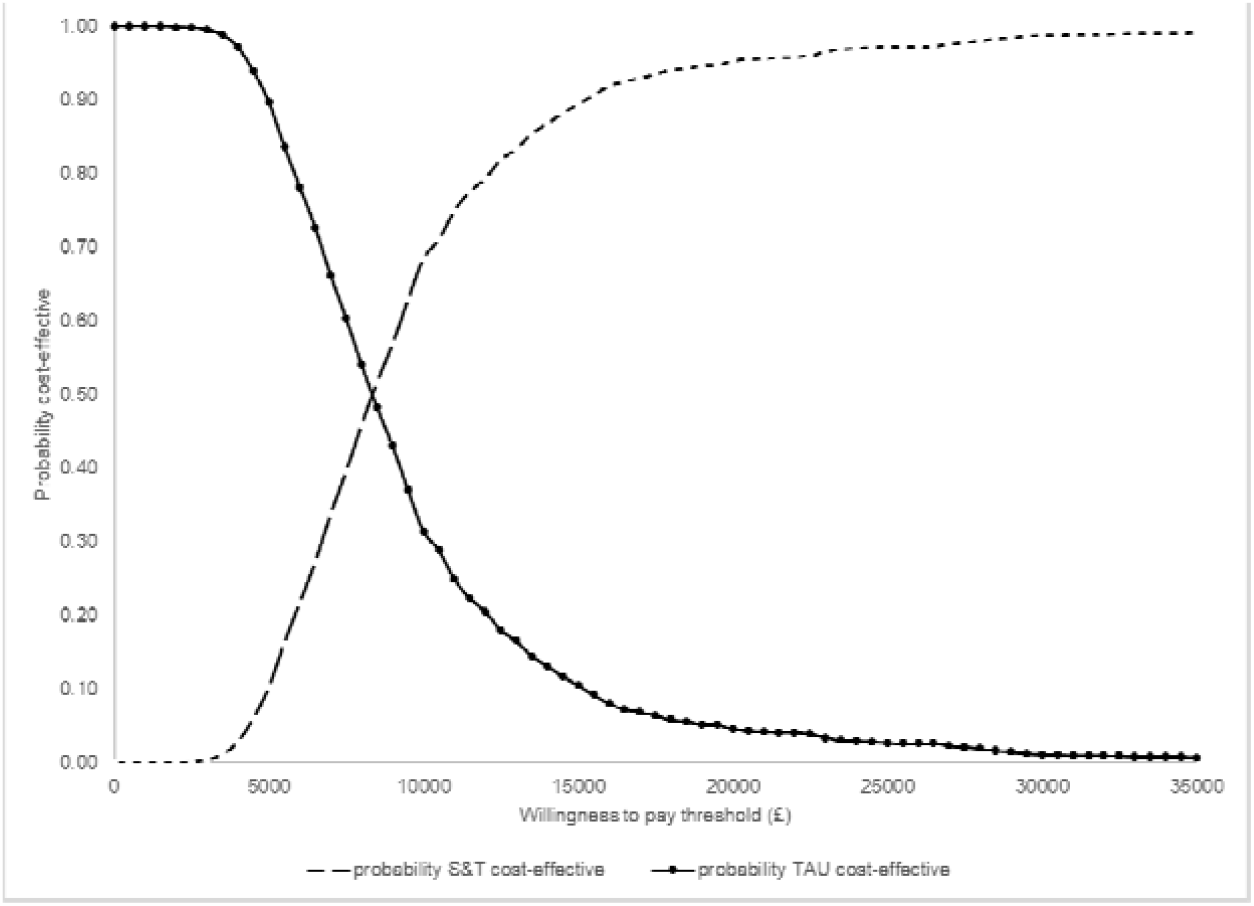
Cost-effectiveness acceptability curve.

## Discussion

The primary aim of the present study was to calculate the incremental cost-effectiveness ratio of a screen-and-treat approach to identifying and treating PTSD in adults following a major incident compared to standard care. It offers important system learning for emergency response planners as the comparator group represents what would likely happen (the counterfactual) if a screen-and-treat approach is not implemented.

Results show an incremental cost-effectiveness ratio of £8,297 per QALY gained. This is below the willingness-to-pay threshold of £20,000 per QALY employed in NICE decision-making contexts and would therefore be seen as representing value for money. This is because, although it costs more than treatment-as-usual, a screen-and-treat approach delivers better population health outcomes that are considered to be large enough to justify the higher costs. Whether this finding warrants utilisation of this approach will ultimately be determined by local decision-makers. Although not directly comparable because of the different target population, CBT for UK middle-aged adults treated for PTSD due to all causes, rather than just for major incidents, was found to be cost-effective, with a net monetary benefit per person over 3 years of £32,042 (Mavranezouli et al., 2020).

The present study has both strengths and limitations. The primary strength is that the economic model was designed to broadly mirror clinical practice. In particular, this was to include multiple rounds of screening and clinical assessment and the changing sensitivity and specificity of the screening tools with time. The model also incorporated heterogeneity in PTSD onset (immediate versus delayed) and recovery following CBT (full versus partial). This study also demonstrated the robustness of the results by addressing uncertainty through deterministic and probabilistic sensitivity analysis. It is a methodological strength that the study has taken a strict NICE perspective because this optimises the relevance of the paper to UK policy-makers.

We limited the economic analysis to a health and social care perspective. This may mean that our estimate of cost-effectiveness is conservative. It is clear from the literature that the economic costs associated with mental health needs, including PTSD, go beyond health and personal social services. For example, indirect costs arising from disrupted employment (Vandentorren et al., 2018) or impaired education (Stene & Dyb, 2019) may be substantial for an individual involved in a major incident. Indeed, empirical analysis of the burden of PTSD following the London bombings found that indirect costs (mostly productivity losses) accounted for the majority (64%) of reported costs (Fuchkan, 2015). Accordingly, the exclusion of indirect costs from the present study may have led to an underestimation of the cost-effectiveness of the screen-and-treat approach.

Another limitation which requires further research is the absence of UK data from a PTSD population in general, let alone within the context of major incidents. Utility values for adults were based on an Australian study for victims of non-terror related trauma (Gospodarevskaya & Segal, 2012). This may not be generalisable to terrorism or major incident-exposed adults in England. However, 0·61 is a conservative approximation for PTSD utility in adults compared with other economic analyses, which potentially reduces the capacity for QALY gains. Another Australian study used reported utility values as low as 0·54 for adults experiencing PTSD (Mihalopoulos et al., 2015). These lower utility values were used in a UK economic modelling study of psychological interventions in a general adult population with PTSD (Mavranezouli et al., 2020). In an economic evaluation of treatment for US military personnel, the QALY value used was also lower with a mean baseline utility value for PTSD of 0.56 (Lavelle et al., 2018).

In assuming that the mental health impact was PTSD, the model did not include depression, anxiety, or substance misuse; this may limit the understanding of cost-effectiveness given the burden of comorbidities. For example, PTSD comorbid with depression has been approximated to have a utility value of 0·53 (Gospodarevskaya & Segal, 2015), which is lower than the 0·61 assumed in the model. In addition, the model did not reflect changes in PTSD prevalence and spontaneous remission rates by age and exposure type (adults, children and first-responders) as it focused on screen-and-treat programmes for adults in the general population exposed to a major incident. This is likely to be the largest sub-population requiring support in the context of the current COVID-19 pandemic.

For first-responders, including health care professionals, a lower prevalence of PTSD would likely decrease the cost-effectiveness of the intervention given the costs associated with the management of false positives. They may be treated more cost-effectively within an occupational health service. Alternatively, following the H1N1 pandemic, research from Japan supported the role of psychiatric liaison services for hospital workers (Matsuishi et al., 2012). For children, any impact on the cost-effectiveness of the intervention of the high prevalence of PTSD may be offset by the minimal QALY differences for children assumed in the literature, namely with PTSD (0.739) compared to without PTSD (0.773) (Shearer et al., 2018). A minimal difference between these utility values, provided this is supported by future research, will reduce the cost-effectiveness of a treatment that transitions a child from having PTSD to not having PTSD.

Our results are potentially generalisable to a variety of emergencies, including pandemics and epidemics. Existing data suggest that certain groups affected by COVID-19, such as survivors of intensive care, will have rates of PTSD that approximate those typical of victims of terrorist attacks (Davydow et al., 2008; Mak et al., 2009; Myhren et al., 2010). Rates of PTSD in frontline hospital staff have also been reported to be high in previous epidemics (Maunder et al., 2006), and can be anticipated to be substantial following COVID-19, especially among care home staff experiencing multiple fatalities, often with a pervasive sense of guilt linked to their perceived role in viral spreading. Furthermore, quarantine measures to control a pandemic have been associated with high rates of PTSD and depression, for example, as observed in a Canadian population quarantined due to SARS (Hawryluck et al., 2004).

It is likely that COVID-19 will also have a profoundly negative impact on psychological outcomes. Indeed, early research from China has shown potentially serious psychological consequences of COVID-19 for healthcare workers and quarantined populations (Chew et al., 2020; Tang et al., 2020). Cost-effectiveness calculations will inevitably be impacted by parameter changes arising from newly acquired knowledge about rates of disorder in different affected groups, and by differences in the ease of identifying and engaging various populations. Some of these changes, such as the increased availability of online screening and treatment, are likely to have the effect of reducing costs (and therefore increasing cost-effectiveness). Others, such as a high prevalence of affected people with a limited knowledge of the host country language, are likely to have the effect of increasing costs. Our model nevertheless provides a structure within which to assess the likely benefits – in the absence of directly observed data at this stage – of instituting screen-and-treat programmes in this different context.

## Conclusion

With the psychological impact of terrorist attacks in recent years still prevalent across many parts of society, the psychological impact of the COVID-19 pandemic is only just beginning to be appreciated. With both forms of major incident, the sequalae of severe psychological conditions, including PTSD, are inevitable features, causing extensive personal and community suffering. Addressing these conditions requires active investment, planning and delivery at all levels from policy makers, strategic and operational health and care teams, and local communities. We hope that the analyses in this paper will go some way to drive a more positive attitude amongst key stakeholders to ensure that services for post-traumatic psychological conditions are funded and managed effectively in this era of major incidents.

## Data Availability

All economic data analysis can be provided on request to the uploading author

## Disclaimers

Neither Patients nor the public was involved in the design, or conduct, or reporting, or dissemination of our findings.

The manuscript is an honest, accurate, and transparent account of the study being reported. No important aspects of the study have been omitted.

## References

Andrews, B., Brewin, C.R, Philpott, R., & Stewart, L. (2007). Delayed onset post-traumatic stress disorder: A systematic review of the evidence. American Journal of Psychiatry, 164(9), 1319–1326. doi:10.1176/appi.ajp.2007.06091491.

Bonanno, G.A., Brewin, C.R., Kaniasty, K. & LaGreca, A.M. (2010). Weighing the costs of disaster: Consequences, risks, and resilience in individuals, families, and communities. Psychological Science in the Public Interest, 11, 1–49.

Brewin, C.R., Rose, S., Andrews, B., Green, J., Tata, P., McEvedy, C., Turner, S.W., & Foa, E.B. (2002). A brief screening instrument for posttraumatic stress disorder. British Journal of Psychiatry, 181, 158–162. doi:10.1017/s0007125000161896.

Brewin, C.R., Scragg, P., Robertson, M., Thompson, M., d‘Ardenne, P., & Ehlers A. (2008). Promoting mental health following the London bombings: A screen and treat approach. Journal of Traumatic Stress, 21(1), 3–8. doi:10.1002/jts.20310.

Brewin, C.R., Fuchkan, N., Huntley, Z., Robertson, M., Thompson, M., Scragg, P., d‘Ardenne, P., & Ehlers, A. (2010a). Outreach and screening following the 2005 London bombings: usage and outcomes. Psychological Medicine, 40, 2049–2057. doi:10.1017/S0033291710000206.

Brewin, C.R, Fuchkan, N., & Huntley, Z. (2010b). Diagnostic accuracy of the trauma screening questionaire after the 2005 London bombings. Journal of Traumatic Stress, 23(3), 393–398. doi:10.1002/jts.20529.

Chew, N., Lee, G., Tan, B., Jing, M., Goh, Y., Ngiam, N., Yeo., L, et al. (2020) A multinational, multicentre study on the psychological outcomes and associated physical symptoms amongst healthcare workers during COVID-19 outbreak [published online ahead of print, 2020 Apr 21]. Brain Behaviour and Immunity. doi:10.1016/j.bbi.2020.04.049.

Cyhlarova, E., Knapp, M., and Mays, N. (2019) Responding to the mental health consequences of the 2015-2016 terrorist attacks in Tunisia, Paris and Brussels: Implementation and treatment experiences in the United Kingdom [published online ahead of print, 2019 Nov 26]. Journal Health Services Research Policy. doi:10.1177/1355819619878756

Curtis, L., & Burns, A. (2018). Unit costs of health and social care. Canterbury: Personal Social Services Research Unit, University of Kent at Canterbury.

Davydow, DS, Desai, SV, Needham, DM, Bienvenu, OJ. (2008). Psychiatric morbidity in survivors of the acute respiratory distress syndrome: A systematic review. Psychosomatic Medicine, 70, 512–519. doi:10.1097/PSY.0b013e31816aa0dd.

Department of Health (DoH), Emergency Preparedness Division. (2009). NHS emergency planning guidance: Planning for the psychological and mental health care of people affected by major incidents and disasters: Interim national strategic guidance. UK. Retrieved from https://webarchive.nationalarchives.gov.uk/20130124050848/ http://www.dh.gov.uk/prod_consum_dh/groups/dh_digitalassets/documents/digitalasset/dh_103563.pdf.

Dyb, G., Jensen, T., Glad, K., Nygaard, E., & Thoresen, S. (2014). Early outreach to survivors of the shootings in Norway on the 22nd of July 2011. European Journal of Psychotraumatology, 5(SUPPL):1–9. doi:10.3402/ejpt.v5.23523.

Ehlers, A., Gene-Cos, N., & Perrin, S. (2009). Low recognition of posttraumatic stress disorder in primary care. London Journal of Primary Care (Abingdon), 2, 36-42. Retrieved from https://www.ncbi.nlm.nih.gov/pmc/articles/PMC3695460/.

First, M., Williams, J., Karg, R., & Spitzer, R. (2015). Structured clinical interview for DMS-5, research version. Arlington, VA: American Psychiatric Association. Retrieved from https://www.appi.org/products/structured-clinical-interview-for-dsm-5-scid-5.

French, P., Barrett, A., Allsopp, K., Williams, R., Brewin, C.R., Hind, D., Sutton, R., Stancombe, J., & Chitsabesan, P. (2019). Psychological screening of adults and young people following the Manchester Arena incident. British Journal of Psychiatry, Open, 5, e85. doi:10.1192/bjo.2019.61.

Fuchkan, N. (2015). Burden of posttraumatic stress disorder – health, social and economic consequences of exposure to the 2005 London bombings. PhD thesis, London School of Economics and Political Sciences. Retrieved from http://etheses.lse.ac.uk/3274/.

Garcia-Vera, M., Sanz, J., & Gutierrez, S. (2016). A systematic review of the literature on posttraumatic stress disorder in victims of terrorist attacks. Psychological Reports, 119(1), 328–359. doi:10.1177/0033294116658243.

Ghebreyesus, TA. (2020). Addressing mental health needs: an integral part of COVID-19 response. World Psychiatry, 19, 129–30. doi:10.1002/wps.20768

Gobin, M., Rubin, J., Albert, I., Beck, A., Danese, A., Greenberg, N., Grey, N., Smith, P., & Oliver, I. (2018). Outcomes of mental health screening for United Kingdom nationals affected by the 2015-2016 terrorist attacks in Tunisia, Paris, and Brussels. Journal of Traumatic Stress, 31(4), 471–479. doi:10.1002/jts.22317.

Gospodarevskaya, E., & Segal. (2012). Cost-utility analysis of different treatments for post-traumatic stress disorder in sexually abused children. Child and Adolescent Psychiatry and Mental Health, 10(6), 15. doi:10.1186/1753-2000-6-15.

Hawryluck, L., Gold, WL., Robinson, S., Pogorski, S., Galea, S., Styra, R. (2004). SARS control and psychological effects of quarantine,,Toronto, Canada. Emergency Infectious Disease,10 (7),1206–1212. doi:10.3201/eid1007.030703.

HM Treasury. (2018). The Green Book – Central Government Guidance on Appraisal and Evaluation. Retrieved from https://assets.publishing.service.gov.uk/government/uploads/system/uploads/attachment_data/file/685903/The_Green_Book.pdf.

Kerslake Report. (2018). An independent review into the preparedness for, and emergency response to, the Manchester arena attack on 22nd May 2017. Retrieved from https://www.jesip.org.uk/uploads/media/Documents%20Products/Kerslake_Report_Manchester_Are.pdf.

Lavelle T, Kommareddi M, Jaycox LH, Belsher B, Freed MC, Engel CC (2018). Cost-Effectiveness of Collaborative Care for Depression and PTSD in Military Personnel. American Journal of Managed Care, 24(2), 91–98.

Mak IWC, Chu CM, Pan PC, et al. (2009). Long-term psychiatric morbidities among SARS survivors. General Hospital Psychiatry, 31(4), 318–26. doi:10.1016/j.genhosppsych.2009.03.001.

Maslow, CB, Caramanica, K, Welch, AE, Stellman, SD, Brackbill, RM, Farfel, MR. (2015). Trajectories of Scores on a Screening Instrument for PTSD Among World Trade Center Rescue, Recovery, and Clean-Up Workers. Journal of Traumatic Stress, 28, 198–205. doi:10.1002/jts.22011

Matsuishi, K., Kawazoe, A., Imai, H., Ito, A., Mouri, K., Kitamura, N., Miyake, K., Mino, K., Isobe, M., Takamiya, S., Hitokoto, H., and Mita, T. (2012) Psychological impact of the pandemic (H1N1) 2009 on general hospital workers in Kobe. Psychiatry and Clinical Neurosciences, 66, 353–360. doi:10.111/j.1440-1819.2012.02336.x.

Maunder RG, Lancee WJ, Balderson KE, et al. (2006). Long-term psychological and occupational effects of providing hospital healthcare during SARS outbreak. Emerging Infectious Diseases, 12(12), 1924–32.

Mavranezouli I., Megnin-Viggars O., Grey N., Bhutani G., Leach J., Daly C., Dias S., Welton NJ., Katona C., El-Leithy S., Greenberg N., Stockton S., and Pilling S. (2020). Cost-effectiveness of psychological treatments for post-traumatic stress disorder in adults. PloS one, 15(4), e0232245. https://doi.org/10.1371/journal.pone.0232245

Myhren, H, Ekeberg, O, Toien, K, Karlsson, S, Stokland, (2010). Posttraumatic stress, anxiety and depression symptoms in patients during the first year post intensive care unit discharge. O. Critical Care, 14. doi:10.1186/cc8870.

Mihalopoulos, C., Magnus, A., Lal, A., Dell, L., Forbes, D., & Phelps, A. (2015). Is implementation of the 2013 Australian treatment guidelines for posttraumatic stress disorder cost-effective compared to current practice? A cost-utility analysis using QALYs and DALYs. Australian and New Zealand Journal of Psychiatry, 49(4), 360–376. doi:10.1177/0004867414553948.

National Institute for Health and Care Excellence (NICE). (2018a). Post-traumatic stress disorder: NICE guidelines [NG116]. Retrieved from https://www.nice.org.uk/guidance/ng116.

National Institute for Health and Care Excellence (NICE). (2018b). Post-traumatic stress disorder: management (update), Evidence Reviews for psychological, psychosocial and other non-pharmacological interventions for the treatment of PTSD in adults [NG116]. Retrieved from https://www.nice.org.uk/guidance/ng116/documents/evidence-review-3.

Neria, Y., Nandi, A. and Galea, S. (2008). Post-traumatic stress disorder following disasters: a systematic review. Psychological Medicine, 38(4), 467–480. doi:10.1017/S0033291707001353.

Pfefferbaum B, North CS, Flynn BW, Norris FH,DeMartino R (2002). Disaster mental health services following the 1995 Oklahoma City bombing: modifying approaches to address terrorism. CNS Spectrums, 7,575–579.

Santiago, P., Ursano, R., Gray, C., Pynoos, R., Spiegel, D., Lewis-Fernandez, R., Friedman, M., & Fullerton, C. (2013). A systematic review of PTSD prevalence and trajectories in DSM-5 defined trauma exposed populations: Intentional and non-intentional traumatic events. PLoS One, 8(4), e59236. doi:10.1371/journalpone0059236.

Shearer, J., Papanikolaou, N., Meiser-Stedman, R., McKinnon, A., Dalgleish, T., Smith, P., Dixon, C., & Byford, S. (2018). Cost-effectiveness of cognitive therapy as an early intervention for post-traumatic stress disorder in children and adolescents: a trial based evaluation and model. The Journal of Child Psychology and Psychiatry, 59(7), 773–780. doi:10.111/jcpp.12851.

Stene, L. and Dyb, G. (2019). Returning to school after a terror attack: a longitudinal study of school functioning and health in terror exposed youth. European Child & Adolescent Psychiatry, 28, 319–328. doi:10.1007/s00787-018-1196-y.

Tang, W., Hu, T., Hu, B., Jin, C., Wang, G., Xie, C., Chen, S., and Xu, J. (2020) Prevalence and correlates of PTSD and depressive symptoms one month after the outbreak of the COVID-19 epidemic in a sample of home-quarantined Chinese university students [published online ahead of print, 2020 May 13]. Journal Affective Disorders. doi:10.1016/j.jad.2020.05.009.

Vandentorren, S., Pirard, P., Sanna, A., Aubert, L., Motreff, Y., Dantchev, N., Lesieur, S., Chavin, P., & Baubet, T. (2018). Healthcare provision and the psychological, somatic and social impact on people involved in the terror attacks in January 2015 in Paris: cohort study. The British Journal of Psychiatry, 212(4), 207–214. doi:10.1192/bjp.2017.63.

Wang, PS; Berglund, P; Olfson, M; et al. (2005). Failure and delay in initial treatment contact after first onset of mental disorders in the national comorbidity survey replication. Archives of General Psychiatry, 62 (6), 603–613.

Whalley, M. & Brewin, C.R. (2007). Mental health following terrorist attacks. The British Journal of Psychiatry, 190, 94–96. doi:10.1192/bjp.bp.106.026427.

WTC Medical Working Group (2008). Annual Report on 9/11 Health. World Trade Center Medical Working Group of New York City: New York.

